# Pilot assessment of low NK cell-mediated ADCC and *FCGR3A* genetics in myalgic encephalomyelitis/chronic fatigue syndrome (ME/CFS): Based on inclusion of family members without ME/CFS as controls, low ADCC is unsuitable as a diagnostic biomarker

**DOI:** 10.1101/2019.12.20.19015438

**Authors:** Alexander P. Sung, Jennifer J-J Tang, Michael J. Guglielmo, Julie Smith-Gagen, Lucinda Bateman, Doug D. Redelman, Dorothy Hudig

**Affiliations:** University of Nevada, Reno School of Medicine, Reno, NV; University of Nevada, Reno Health Sciences; Bateman Horne Center, Salt Lake City, UT; Deceased, University of Nevada, Reno School of Medicine, Reno, NV

**Keywords:** Myalgic Encephalomyelitis, Chronic Fatigue Syndrome, ADCC, antibody-dependent cell-mediated cytotoxicity, NK cell, CD16A, family studies, biomarker

## Abstract

ADCC (antibody-dependent cell-mediated cytotoxicity) is dependent on the varying capacity of NK cells to kill, the affinities of *FCGR3A*-encoded CD16A receptors for antibody, and the presence of antigen-specific antibodies. *In vivo* ADCC depends on the number of CD16A receptor-positive NK cells in blood. We hypothesized that low ADCC cell function or low effector cell numbers could be biomarkers or risk factors for myalgic encephalomyelitis/chronic fatigue syndrome (ME/CFS). We measured NK cell ADCC lytic capacity and antibody recognition, CD16Apositive NK cells/µl blood, and *FCGR3A* homozygosity for the F allele that encodes low affinity CD16A antibody receptors. ME/CFS patients met the Fukuda 1994 diagnostic criteria. In this pilot report, we examined 5 families, each with 2 to 5 ME/CFS patients, and compared 11 patients, 22 family members without ME/CFS, and 16 unrelated healthy controls. ADCC was measured as CX1:1 cytotoxic capacity (the percentage of ^51^Cr-Daudi tumors with obinutuzumab anti-CD20 antibody that were killed at a 1:1 ratio of CD16Apos NKs to Daudis) and CX-slope. Individual CX1:1 capacities varied from 16.2% to 81.8% and were comparable between patients and unaffected family members, while the ADCC of both family groups was lower than the unrelated healthy controls. The lack of difference between patients and their unaffected family members indicates that low ADCC is unsuitable as a diagnostic biomarker for ME/CFS. Familial CD16Apos NK blood cell counts were lower than unrelated healthy controls. The potential for synergistic effects of combined low CX1:1 and low effector cell counts occurring in the same individual was 24-fold greater for CFS family members than for unrelated controls. *FCGR3A* of the families was predominantly F/F homozygous, correlating with the observed low EC50 for NK recognition of target cell-bound antibody. In summary, low ADCC is unsuitable as a biomarker, but could be a familial risk factor, for ME/CFS.

## Introduction

Myalgic encephalomyelitis/chronic fatigue syndrome (ME/CFS) is an umbrella term, with ME and CFS often used interchangeably, that is used to describe patients who meet the evidence-based diagnostic criteria described by the National Academy of Medicine USA in 2015 [1]. The National Academy suggested an alternate term, systemic exertion intolerance disease (SEID) that has yet to be incorporated into the scientific or popular literature. The medical condition lacks unique diagnostic biomarkers and characterization of the underlying mechanisms of its pathology. ME/CFS has common debilitating symptoms that affect as many as 800,000 to 2.5 million adults in the USA [1]. These symptoms include severe fatigue lasting more than six months, long-lasting post-exertional malaise, un-refreshing sleep, ‘brain fog’ in the form of loss of memory or lessened ability to think, chronic pain, and tender lymph nodes [2]. There are no known causes for most cases; however, long-term ME/CFS-like pathology can follow severe viral or bacterial infections [3]. ME/CFS is receiving increasing attention as a distinct disease [4] and is viewed by some clinicians as being as devastating as HIV/AIDS [5]. ME/CFS is quite costly to society [6]. Afflicted individuals would benefit from greater understanding of its etiology and disease-promoting mechanisms and from discovery of definitive diagnostic biomarkers. To the best of our knowledge, this pilot report is the first study since 1998 [7] that compares biological functions of ME/CFS patients *vs*. their first degree family members in the search for biomarkers. The inclusion of family members without ME/CFS as controls in the present report resulted in the rapid identification of unsuitable diagnostic ADCC biomarkers (which would have “passed” evaluation if comparisons were made only of patients *vs*. the unrelated healthy controls).

Consideration of infectious and environmental etiologies (that could evoke ADCC) began when the disease, which was originally termed as CFS, was first recognized in 1984 in a regional cluster of patients [8]. Several viruses have been proposed to exacerbate CFS, including chronic herpes viruses such as Epstein Barr virus [9-11], human cytomegalovirus (HCMV) [12], herpes zoster [13], human herpes virus 6 (HHV6) [14-16], and parvovirus B19 [17, 18]. The fatigue of CFS resembles the fatigue induced by gamma interferon during viral infections [19, 20]. Elevated gamma interferon is detectable in the blood of patients with severe CFS [21]. However, attempts to prove that viral infections are associated with most cases of CFS have been generally unsuccessful [22, 23]. The potential for genetic predisposition to contribute to etiology was first reported in 1998 by P.H. Levine *et al*. [7] for eight CFS patients within one family. Monozygotic twins have reported concordance rates as high as 38% for a USA population of female twins [24]. Other reports indicated lower concordance rates or little difference in Swedish twins [25] and in a USA twin registry [26]. Attempts to identify a unique viral etiology or detect unique gene expression in the CFS discordant monozygotic twin pairs [27] have been unyielding to date. To date, both infectious causes and familial risks are unresolved issues in CFS.

NK cells mediate both nonspecific anti-viral immunity and specific anti-viral antibody-dependent cell-mediated cytotoxicity (ADCC) when anti-viral antibodies are present. Caligiuri *et al*. observed in 1987 that CFS patients have low NK activity [28]. Studies followed that reported low NK function (reviewed [29], also [30-33]), NK phenotypes consistent with lower NK cell function (*e.g*., low perforin [34]; low granzyme B [33]; lower frequency of the more cytotoxic CD56dim NK cells [35], and low ERK1/2 kinase [36]), and fewer blood NK cells in patients during CFS disease episodes [37]. In contrast, a recent Scandinavian study with large numbers of ME/CFS patients indicates that many CFS patients have normal NK activity and that low NK activity is unsuitable as a biomarker for this disease [38]. Consequently, low NK cell activities in ME/CFS must be viewed as far from universal.

We began this study with the idea that low ADCC mediated by NK cells could be a risk factor for CFS, the term we use for identification of its patients. This ADCC is dependent on only those NK cells that have CD16A receptors for antibodies and on the fraction of these CD16A-positive NK cells that can kill. Even though the NK cells that mediate ADCC and natural cytotoxicity use the same cytotoxic granule proteins, it is possible for natural cytotoxicity to be compromised without affecting ADCC. For example, a rare mutation of the cytoplasmic region of CD16A (where it binds to CD2) reduces natural cytotoxicity to K562 cells without affecting antibody-recognition and ADCC [39, 40]. Also, variants of NK receptors [40] which are involved in natural cytotoxicity (but unnecessary for ADCC) will affect the NK activity without affecting ADCC. Thus, when natural cytotoxicity is reduced in CFS patients, ADCC could remain unaltered and, when ADCC is affected, natural cytotoxicity could remain unaltered. We hypothesized that ADCC risk factors for CFS might include: 1) reduced ADCC cellular capacities, 2) decreased ADCC effector cell numbers and/or 3) requirement for more antibody to support ADCC. Specifically, here we addressed: a) NK cell cytotoxic capacity for ADCC; b) counts of blood NK cells with CD16A receptors; c) homozygosity for the *FCGR3A* allele that encodes the IgG-Fc-receptor CD16A aa158 phenylalanine (F) that has lower affinity for IgG1 antibodies than aa158 valine (V) [41]; and d) the EC50 concentrations of antibody needed for NK cells to recognize antibody-coated cells. The numbers of NK cells in blood are particularly important because the NK cells in blood are responsible for most of human ADCC while most tissue NK cells lack CD16A and ADCC [42, 43].

The study reported here has several novel features, including the ADCC measurements used for comparisons and the inclusion of family members without ME/CFS as a second set of controls. Two methods were developed for the study: an ADCC assay to detect cytotoxic capacity independent of *FCGR3A* Fc-receptor genotypes [44] and TruCount® bead enumeration of CD16A-positive(pos) blood NK cells. The ADCC assay uses a type 2 anti-CD20 monoclonal antibody (obinutuzumab) to pretreat CD20pos B cell tumor ‘target’ cells, so that the normal B cells within PBMCs are unable to serve as ADCC targets and assays can be made without the cell losses associated with NK cell purification. The CD16Apos NK cells within unfractionated PBMCs were quantified by flow cytometry (with TruCount® beads) to determine the effector to target ratios in the ADCC ^51^Cr-release assays. To the best of our knowledge, this report is the first to address ADCC functions in CFS patients compared to their family members. The report focuses on members of five families, each with several CFS patients. Shared overall genetic backgrounds and early environmental conditions could promote detection of specific alleles and non-genetic adaptive changes in NK cells [45] that might affect CFS. Comparison of affected *vs*. non-affected family members has the potential to identify rigorous diagnostic biomarkers.

We report five observations. (1) There was lower ADCC capacity by NK cells of CFS patients compared to unrelated healthy controls. (2) There was no difference in ADCC between CFS patients and their family members without CFS, *clearly indicating that low ADCC is unsuitable as a specific biomarker for CFS*. (3) ADCC blood CD16Apos NK cell counts of CFS patients and their family members without CFS were lower than the counts of unrelated healthy controls.(4) CFS family members (with or without the disease) were much more likely to have a combination of low ADCC activity with low CD16A NK cell counts than the unrelated healthy controls. (5) The CFS families were predominantly homozygous CD16A F/F. On the basis of these observations, we suggest that, while low cellular ADCC activity *alone* is definitely not a biomarker of CFS, low ADCC may be a risk factor for CFS.

## Materials and Methods

### CFS patients, family members and unrelated healthy donors

Five families, available at the Bateman Horne Center in Salt Lake City, UT, each with 2 to 5 CFS patients, were selected by Dr. Lucinda Bateman and the research team from many families afflicted with CFS. The majority of these CFS patients had myalgia; their disease is designated here simply as CFS because they were diagnosed by the Fukuda criteria during the period between 1990 and 2010, before the terms ME or ME/CFS became common in the U.S.A. Selection was for families each with several CFS patients and with multiple unaffected siblings. The initial idea was to compare the incidence of CFS among CD16A F/F *vs*. V/V & V/F siblings, with the hypothesis that the CFS incidence would be greater for the F/F siblings. (This hypothesis remains untested as almost all family members of this study were F/F.) The five families selected had a total of 13 CFS patients and three families had two generations with CFS patients. The CFS patients met the Fukuda CDC 1994 diagnostic criteria for CFS [2]. Eleven of the 13 CFS patients and 22 family members without CFS participated. The mean ages, sex, and CD16A genotypes of the participating patients, their unaffected family members and the controls are presented in Table 1. The ages, symptoms that qualify for the Fukuda criteria, and durations of CFS disease of the patients are detailed in Table 2. Of the 11 CFS patients, 9 were female and 2 male, representative of the gender bias of CFS. The family pedigrees are illustrated in Fig 1; participants in the study have their CD16A aa158 alleles within the symbols. Selection was based on family members’ willingness to participate and their availability to donate blood on the same day as did the patients. Additional information concerning ages, symptoms, duration of disease, *etc*. of the patients may be obtained from Dr. Bateman upon request. Sixteen unrelated healthy control donors who resided in Salt Lake City were matched by race, sex and age to the patients. Healthy was defined as HIV-negative, without overt infections at the time of blood donation, and without diagnosis of CFS. All blood donors, family and unrelated, were Caucasian. Blood samples were coded in Salt Lake City. The ADCC, EC50 assays, blood CD16A NK cell counts, and CD16A genotypes were run with the coded samples and decoded after completion of the experiments. Most shipments of blood samples included multiple CFS family members and two unrelated healthy controls. The research with human subjects was approved by institutional review boards (IRBs) for the Bateman Horne Center and for the University of Nevada, Reno School of Medicine. Written informed consent was obtained from the blood donors.

**Table 1.** Characterisistics of the Three Study Populations

| Table 1. Characteristics of the Three Study Populations |  |  |  |  |  |  |
| --- | --- | --- | --- | --- | --- | --- |
| Characteristic | CFS Cases |  | Family members w/o CFS |  | Unrelated healthy controls |  |
| Number of participants | 11 |  | 22 |  | 16 |  |
| Number of families | 5 |  | 5 |  | 16 |  |
| Age, mean +/- SD | 45.4 +/- 21.3 |  | 46.2 +/- 14.7 |  | 42.8 +/- 18.6 |  |
|  | Number | % | Number | % | Number | % |
| Sex |  |  |  |  |  |  |
| Female | 9 | 81.8 | 13 | 59.1 | 12 | 75.0 |
| Male | 2 | 18.2 | 9 | 40.9 | 4 | 25.0 |
| CD16A AA158 genotype |  |  |  |  |  |  |
| F/F | 10 | 90.9 | 18 | 81.8 | 3 | 18.8 |
| F/V or V/V | 1 | 9.1 | 4 | 18.2 | 13 | 81.3 |

**Table 2.**
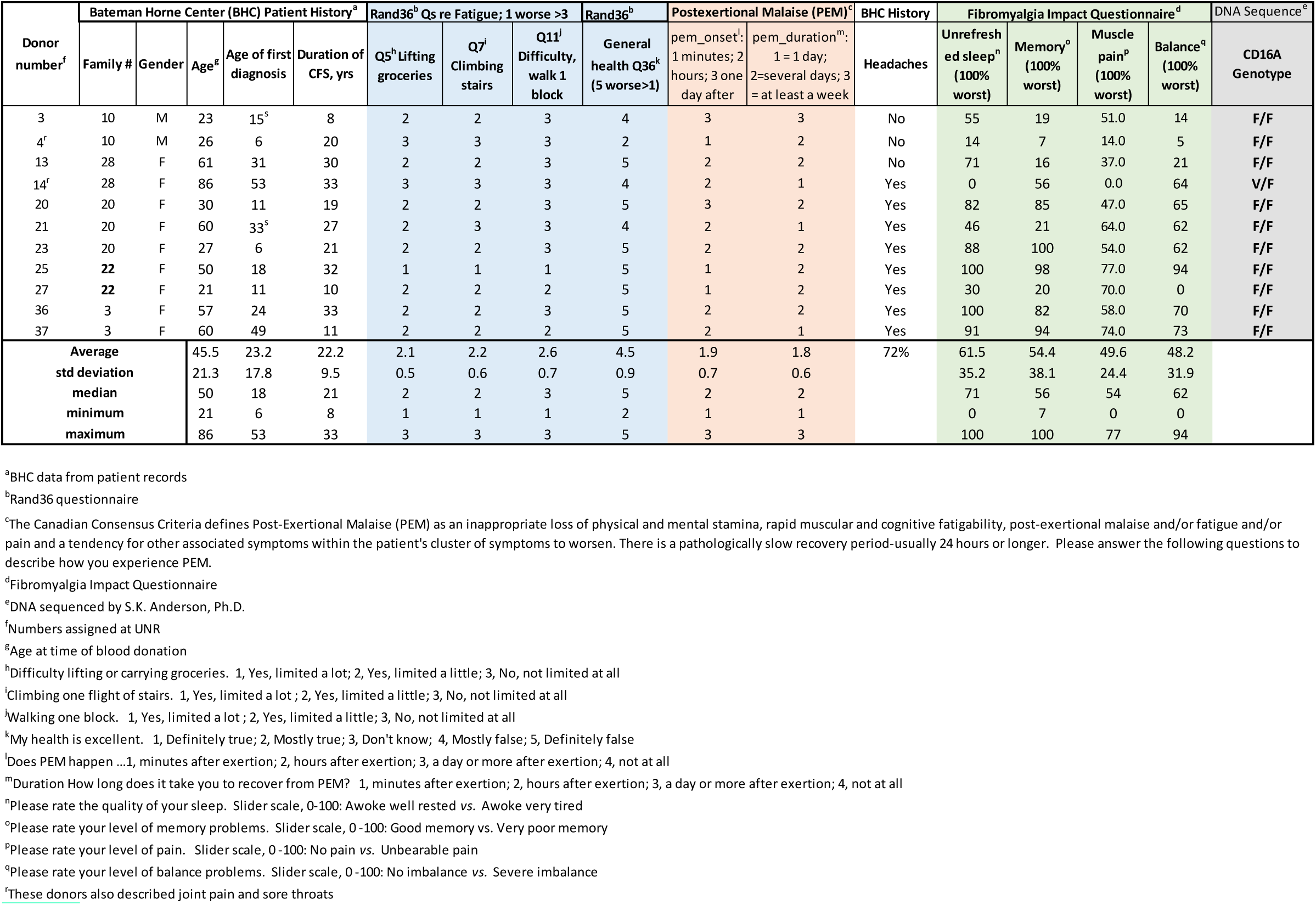
Characteristics of the ME/CFS Patients

**Fig 1.**
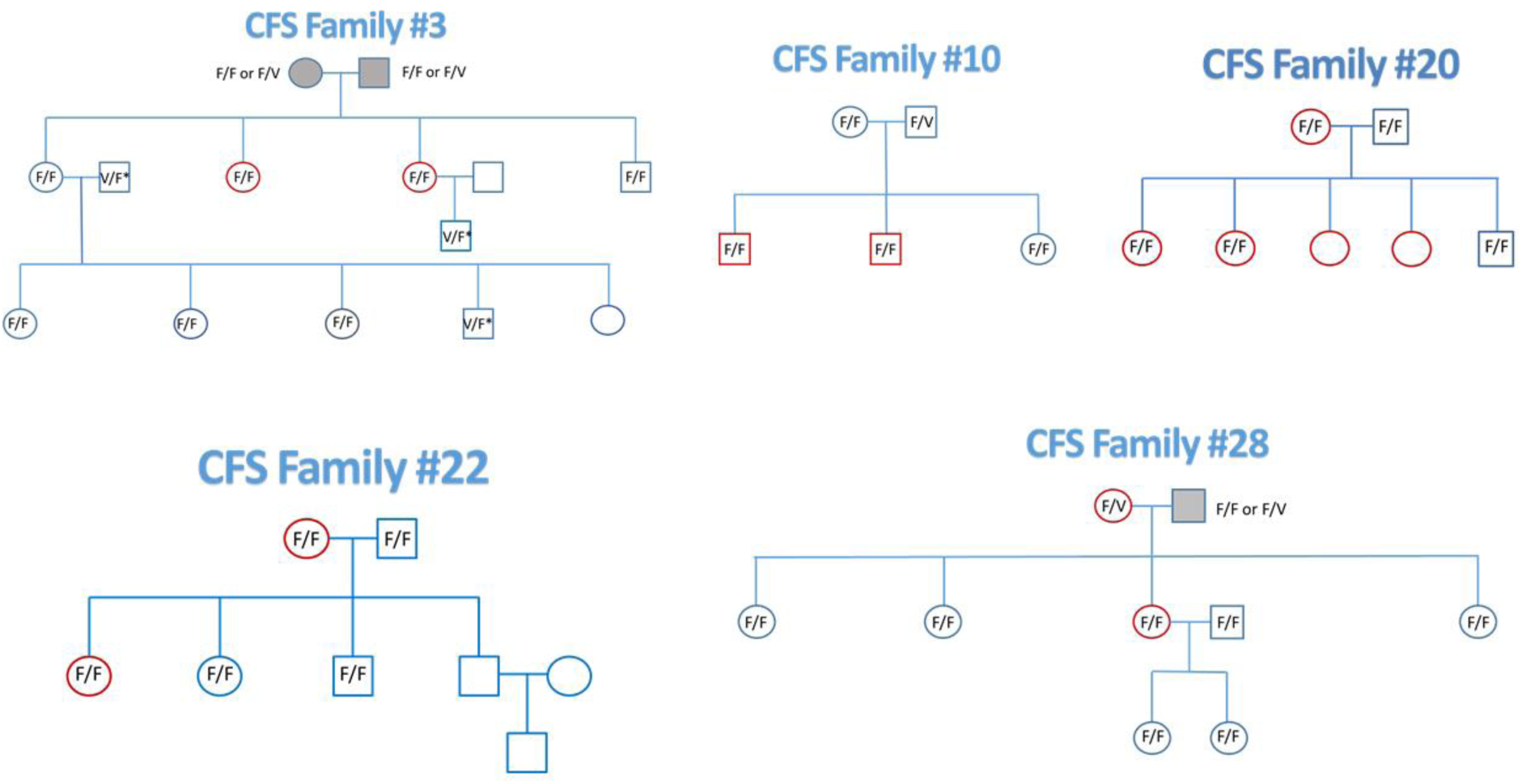
CFS Family pedigrees & CD16A genotypes. The pedigrees of the 5 CFS families are illustrated. Red symbols indicate CFS patients. The participants in the study are indicated by their CD16A aa158 alleles. Family members who were unavailable are indicated by open symbols without CD16A alleles. The 3 deceased parents are indicated with gray symbols. The CD16A genotypes were determined by DNA sequencing and by flow cytometry. Three genotypes in family 3 (marked V/F* in the pedigree) were determined only by flow cytometry (which detects CD16A aa158 V) in combination with pedigree analysis of the (sequenced) alleles that these individuals transferred to their progeny.

### Preparation of peripheral blood mononuclear cells (PBMCs)

Forty ml of blood was drawn from the subjects in Salt Lake City, between 8-10 AM and distributed as follows; 8 ml into tubes for DNA extraction (PAXgene®, Qiagen, a BD company) and the remainder 32 ml into heparinized tubes. Entire families, or groups of members of the larger families, with inclusion of 1 or 2 unrelated healthy controls, were drawn on the same day. The samples were coded and shipped overnight to Reno, NV. PBMCs from 24 ml of blood were isolated at UNR by ficoll-hypaque density gradient centrifugation [46]. The PBMCs were cultured overnight at 1-2 × 10^6^ cells/ml in complete assay media, 90% Dulbecco’ s complete media containing high (4.5 g/L) glucose and L-glutamine (Corning), 10% fetal calf serum (Atlanta Biologicals), 10 mM hepes (Sigma-Aldrich, St. Louis, MO), and 1% penicillin-streptomycin (Sigma-Aldrich) to ensure ADCC activity [47]. Culture conditions and assay media were standardized: one lot of fetal calf serum was frozen and used for all the experiments and one lot of 75 mm tissue culture flasks (Biolite, Thermo Scientific) was used throughout the experiments.

### TruCounts® of CD16A NK effector cells

#### NK effector cells per µl blood

Fifty µl aliquots of whole blood were labeled on the days of arrival with the following panel of antibodies designed for no-wash analyses with TruCount® beads: PacBlue anti-**CD45** (clone HI30); FITC-anti-**CD3e** (clone UCHT1); FITC-anti-**CD91** (2MR-alpha); and PerCP-anti-**CD16A** (3G8); purchased from BioLegend (San Diego, CA) with the exception of the anti-CD91 from Becton Dickenson. The NK ADCC effector cells were CD3negCD16posCD45pos & CD91neg.

#### NK effector cells per µl PBMC solution

For the counts of the NK cells within the PBMCS, 50 µl aliquots of PBMCs were labeled with a similar panel of antibodies, with the substitution of PE-Cy7 anti-**CD33** (clone P67.6) for CD91 to detect monocytes and inclusion of FITC-anti-**CD7** (clone CD7-6B7) and APC-Cy7 anti-**CD56** (clone HCD56). The NK ADCC effector cells were CD3neg CD7posCD16pos CD33negCD45pos and CD56variable. Inclusion of anti-CD7 was critical to detect CD56negCD16AposCD7pos NK cells and to discriminate these NK cells [48] from the CD56negCD16Apos monocytes (that are all CD7neg and largely CD33pos) [49].

#### Cytometry to detect NK effector cells

Cells were labeled for 30 minutes in tubes with TruCount® beads (Becton Dickenson no. 340334 [50]), fixed, and analyzed by flow cytometry the same day. The flow cytometer was a BD Biosciences Special Order Research Product LSR II analytical flow cytometer with a high throughput sampler (HTS) unit. Cellular analyses were gated with FlowJo software (FlowJo, LLC, Ashland, OR) to determine the CD16Apos NK cells and TruCount® beads in order to calculate the number of CD16Apos NK cells in the PBMC solutions [44]. See Fig 4B for the cytometric gating.

**Fig 2.**
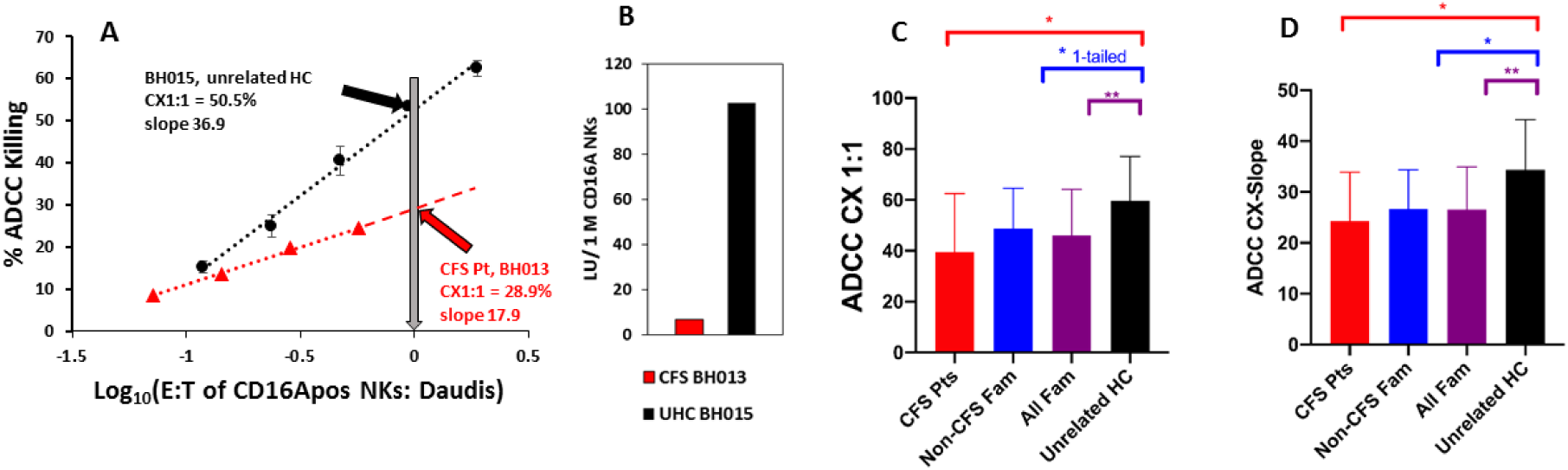
ADCC of CFS patients, family members without CFS, and unrelated controls. CFS family members, both diagnosed and without CFS, had lower ADCC than unrelated healthy donors. **A&B. Representative ADCC methodology for a CFS patient and a healthy unrelated donor**. The ADCC assays illustrated were performed on the same day. **A. CX1:1 and CX-slope**. CX1:1 values, the percentages of target cells killed at a 1 to1 ratio of CD16A (IgG FcR)-positive NK cells to Daudi targets, are indicated by the arrows. The CX-slopes are indicated by text in the graph. The error bars indicate standard deviations for 4 points per effector to target (E:T) ratio. The P values of the linear fits were <0.001 and the R^2^s >0.96. The NK activity towards Daudi cells (*without* antibody) was below 5% for both donors at their highest E:T ratios and is not illustrated. **B. Lytic units**. Lytic units (LU_50_) of ADCC per 10^6^ CD16Apos NK cells for the two donors of Fig 2A are presented to indicate how the nonparallel and lower CX-slope of the CFS patient amplifies differences when lytic units are used for comparisons. **C&D. Comparisons of ADCC**. Red bars indicate CFS patients, blue bars indicate their family members without CFS, purple bars indicate all family members combined, and black bars indicate unrelated healthy donors. Mean values with standard deviations are indicated. Single asterisks above the brackets indicate Ps<0.05, two asterisks Ps <0.01, for two-tailed T-test comparisons with the unrelated controls. **C. CX1:1 was lower for CFS patients and for all CFS family members compared to the unrelated healthy donors**. CFS patients and their family members without disease had similar CX1:1s, both of which were lower than the unrelated healthy controls. The average CX1:1 values of CFS, healthy family, and unrelated healthy donors was 39.5%, 47.8% and 59.7%, respectively. The average CX1:1 of all family members was 45.1%, 77% of the CX1:1 of the unrelated healthy controls. The CX1:1 data are from families #3, 10 & 28 and additional data for these donors are illustrated in Figs 4A and 5A.**D**. T**he CX-slopes, indicative of cellular cooperativity in killing, were lower for the CFS patients and for all CFS family members compared to the unrelated healthy donors**. CX-slopes are the % increase in dead cells per 10-fold increase in effector cells. The color codes are the same as in **C**. The average CX-slope values of CFS, healthy family, and unrelated healthy donors were 24.3%, 25.2% and 34.4%, respectively. The average CX-slopes of all CFS family members combined was 24.8%, 75% of that of the unrelated healthy controls. CX-slope data are from families #3, 10, 20, 22 & 28.

**Fig 3.**
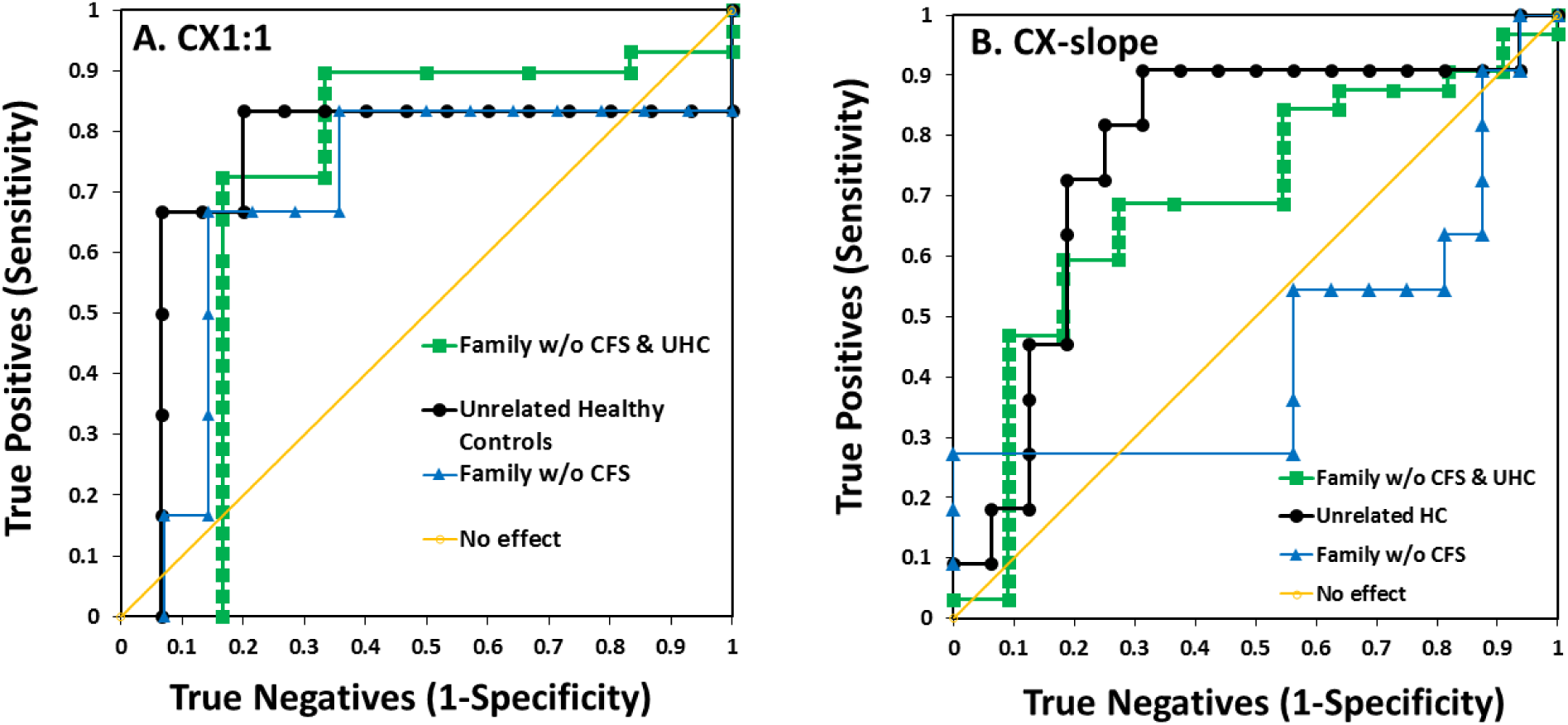
ROC area under the curve (AUC) analyses of ADCC. ROC AUCs indicate unsuitability of low ADCC as a biomarker to diagnose CFS, particularly when applied to the unaffected family members of CFS patients. ROC analyses are illustrated for (**A**) **CX1:1** and (**B**) **CX-slope**, using the data illustrated in Fig 2. The ‘true positives’ of the ROCs are the CFS patients compared with ‘true negatives’ of: 1) only the unrelated healthy controls; 2) all donors in the study without CFS, including the unrelated healthy donors and the family members without CFS; and 3) only family members without CFS. The diagonal line of 0.5 AUC illustrates values for tests with no difference between true positives and true negatives.

**Fig 4.**
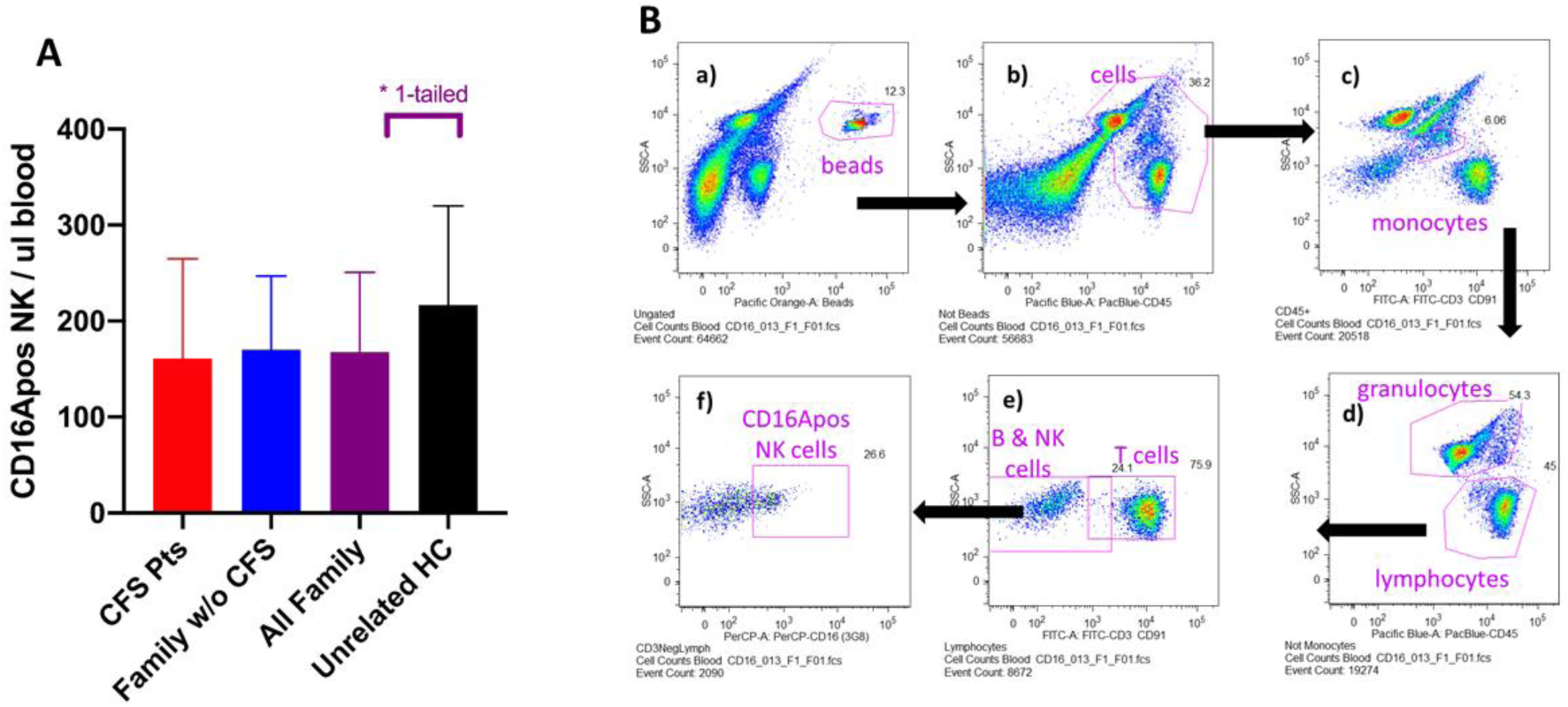
NK effector cell counts. CD16Apos NK effector cell counts per µl of blood as determined with TruCount^R^ bead calibration of the blood volumes sampled. **A. Counts of NK cells with CD16A receptors per µl of peripheral blood**. Symbols use the same color codes as in Figs 2C & 2D. CFS patients and their family members without CFS had lower CD16Apos NK cell counts than the unrelated healthy controls. The average number of cells per µl blood for all CFS family members was 77% of the healthy controls. A one-tailed t-test to determine if CFS family members had lower counts than controls had a statistical significance of P<0.05. Data are from families #3, 10, 22 & 28 and include the donors illustrated in Figs2C and 2D and 5A and 5B. **B. Flow cytometric gating for the TruCount**®**s of CD16Apos NK cells in blood**. The gating for the CD3negCD91negCD16Apos NK cells is illustrated. Letters indicate the sequence for each gate; the arrows indicate the cellular subsets selected for each subsequent gate. Whole blood was placed in TruCount® tubes, a panel of labeled antibodies was added, and the cells and beads were counted *without* washing. FITC anti-CD3 and anti-CD91 mAb’s were used for multiplex staining of T cells and monocytes, respectively. The monocytes could be distinguished by their side scatter. The TruCount® beads are gated in **a)**. The “not beads” events were gated for Pac-blue CD45positive cells in **b)**. The monocytes within the CD45-pos cells were gated by side scatter and FITC anti-CD91 labeling in **c)**. The “not-monocytes” were then assessed to gate out granulocytes by high side scatter in **d)**. The lymphocytes were then assessed for FITC-CD3 to discriminate T cells in **e)**. The remaining B, NK and other cells were next gated to detect CD16Apos NK cells in **f)**. The blood count of CD16Apos NK cells was 69 cells per µl blood for the CFS patient BH013 illustrated, the same patient characterized for NK activity in Fig 2A

### ADCC assays

#### NK cell capacity and cooperativity

The assay measures ADCC calculated for 1:1 ratios of CD16Apos effector NK cells to antibody-coated target cells [44] in order to compare donors with widely varied lytic capacities. It measures the lytic activity of cells and is unaffected by CD16A V and F allelic differences. The assay, which was developed for this study, has four critical features: 1) MHC class I-negative Daudi target cells in order to avoid inter-donor variations caused by KIR inhibition after engagement with MHC I proteins; 2) a type 2 anti-CD20 monoclonal antibody that is poorly cleared from the membranes of B cells so that Daudi B cells could be pretreated with antibodies and washed (thereby preventing the ADCC towards native B cells within the PBMCs that would occur if unbound anti-CD20 antibody were present during the assays); 3) circumvention of NK cell losses during isolation by using TruCount®s of the CD16A receptor-positive NK cells within the PBMCs to determine the effector to target cell ratios; and 4) separate measurement of the lytic slopes because the slopes of killing at multiple effector to target (E:T) ratios vary among donors. The Daudi lymphoma target cell line [51] from the ATCC (Manassas, VA, catalog # CCL-213) was maintained per ATCC instructions and routinely tested negative for mycoplasma. Passages 10-80 were used as targets cells at 1 × 10^4^ cells/well. The non-fucosylated anti-CD20 monoclonal antibody obinutuzumab (Gazyva®; also reported as GA101) [52-54] was obtained from the Roche Innovation Center, Zurich, Switzerland. Daudi target cells were labeled with 0.5 mCi Na^51^CrO_4_ (Perkin Elmer, Waltham, MA), then pretreated with antigen-saturating (1 µg/ml) obinutuzumab antibody for 0.5 hour at room temperature, and washed 5 times to remove excess antibody and unincorporated chromium. The PBMC effector cell solutions were diluted 2-fold in quadruplicate in V-bottom plates (Costar 3894, 96 well) in 0.1 ml to create six CD16Apos NK effector to target cell (E:T) ratios. Daudi cells (1 × 10^4^ in 0.1 ml, with or without mAb pretreatment) were added to each well, for ADCC or NK background activities, respectively. Plates were centrifuged for 3 minutes at 1000 rpm and incubated for 4 hours at 5% CO_2_ and 37°C. After incubation, plates were centrifuged for 10 minutes at 1200 rpm and 0.1 ml of cell-free supernatant was taken for analysis in a Packard Cobre II gamma counter. The percent specific release (SR) was calculated using the formula

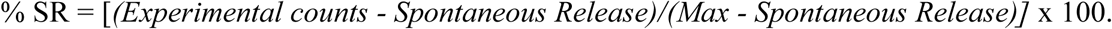

Spontaneous release was the leak rate of targets without PBMCs and the Max was the radioactivity released by targets lysed with 1% SDS. CX1:1 cytotoxic capacity, the % of the targets killed at a 1:1 ratio of effector CD16Apos NK cells to Daudi target cells (E:T), was calculated as follows. Percent ADCC values for individual wells were plotted as y = % specific ^51^Cr release *vs*. x = the log_10_ of the TruCount® NK effector cell to Daudi target cell ratio. The linear % cytotoxicity *vs*. log_10_ E:T was used to calculate y = mx + b, with the lytic slope = m, and b = the y intercept = the CX1:1 value (because for x = 0 the log_10_ E:T is 0 and 10^0^ is 1). The P values for linearity were <0.05, with R^2^ values >0.8 [44]. Lytic units (LUs) per 10^6^ CD16Apos NK cells were calculated [55] to provide reference to previous reports; however, for inter-donor comparisons of LUs to be quantitative, the CX-slopes must be parallel and, in most cases, these CX-slopes differed substantially among donors.

### EC50 assay for NK cells’ antibody recognition

The EC50 effective concentrations of antibody needed for 50% of maximal ADCC [56] were determined with a single concentration of PBMCs and 4-fold dilutions of obinutuzumab in the assays. Final antibody concentrations ranged from 0.04 to 625 ng/ml. ADCC was determined at 4 hours, with duplicate or triplicate wells for each antibody concentration. The yields of PBMCs after ficoll preparation were sufficient to support EC50 determinations for most, but not all, donors.

### Genotyping of *FCGR3A* alleles encoding CD16A F and V variants

#### Genotyping by DNA sequences

The F/F, V/F and V/V genotypes of the receptor CD16A at aa158 were determined by PCR and DNA sequence analysis at the Frederick National Laboratory for Cancer Research, Frederick, MD, by Stephen K. Anderson, Ph.D. and additionally by flow cytometry. Amplicons specific for the *FCGR3A* gene (which encodes CD16A) and that exclude the *FCGR3B* gene (which encodes neutrophil CD16B) were generated with forward and reverse PCR primers, (5’ to 3’) for CD16 (TCCTACTTCTGCAGGGGGCTTGT) and (CCAACTCAACTTCCCAGTGTGATTG), respectively. The amplicons were directly sequenced using Sanger methodology to determine the genotypes [57].

#### Genotyping by flow cytometry

The homozygous F/F genotype was also distinguished from V/F & V/V genotypes by flow cytometry [44] using the MEM-154 clone of anti-CD16 mAb (PE-labeled, Pierce Chemical Co, Rockford, IL) to identify CD16A F/F donors. MEM154 reacts with the CD16A 158 V but not the 158 F [58], and also reacts with CD16B (that has only the valine form). An aliquot of the PBMCs was labeled with the following antibody panel (for gating refer to the legend of supplement Fig 3): FITC-anti-**CD3e** (cloneOKT3); PE-anti-**CD16A** (3G8) or PE anti-**CD16A 158V selective**-(MEM154); BV605-anti-**CD19** (HIB19.11); PacBlue anti-**CD45** (HI30); FITC-anti-**CD91** (2MR-alpha) and APC-Cy7-anti-**CD56** (HCD56), purchased from BioLegend (San Diego, CA) with the exception of the MEM154 mAb. The cells were fixed and washed prior to flow cytometric analyses.

### Efforts to limit intra-experimental variation

The experiments were conducted between November 19, 2015 and January 26, 2017. Inclusion of family members, CFS patients and unrelated healthy controls in a single experiment with 4-6 donors was one means to limit the impact of inter-experimental variations. We also used a single lot of obinutuzumab and the same lots of fluor-tagged antibodies to promote consistency.

### Statistical analyses

The ADCC measurements and linear correlations were determined with the Excel Analysis Tool Pack, using best fit for linear regressions to determine CX1:1 and CX-slope. Student’s t-tests in Excel were used to compare the different groups of subjects. Excel and GraphPad Prism 7 (San Diego, CA) were used for illustrations and calculations of t-tests. The UCSF website http://www.sample-size.net/sample-size-proportions/ was used to determine the 95% confidence limits for the F/F genotypes [calculation: “CI for Proportion”] and to estimate the minimal CFS parental and control sample sizes [calculation: “Proportions—Sample Size”] required to apply the observed F/F frequencies of this study to the design of a larger future study. For the quadrant analyses to examine disease-promoting synergy when two variables coincided (in CFS patients or CFS family members compared to healthy controls), variables were assessed with quadrants delineated by their median values as the cutoffs for low and high values. The three variables analyzed were CX1:1, CX-slope and CD16Apos NK cell counts per µl blood. Two quadrant analyses were synergistic: comparison of CX1:1 & CD16A NK counts and CX-slope & CD16Apos NK counts. Logistic regression using interaction terms was applied for the synergistic comparisons. The results were expressed as the odds of being in the low-low quadrant for CFS patients and family members *vs*. unrelated healthy control subjects. SAS statistical software (SAS v. 9.2 Institute Inc., Cary, NC, USA) was used for the quadrant analysis. Biomarker potentials were assessed using receiver operating characteristics (ROC) analyses [59] with SAS software version 9.4. The area under the curve (AUC) offers a means to compare the value of a biomarker when applied to different groups of patients; a value of 0.5 indicates no difference between the two groups while a value of 1.0 indicates a perfect biomarker test that identifies all and only patients with CFS. The P values for the two estimates of the frequencies of F/F parents were determined using the Appendix to Chapter 5: Exact Binomial Probability Calculator available at http://vassarstats.net/textbook/ch5apx.html. Unavailability of the flow cytometer impacted data collection for a few donors. As a result, there are more measurements of CX-slopes than CX1:1, as the CX1:1s are dependent on flow cytometric TruCount®s. The data for individual members within each of the 5 families and the healthy controls are illustrated in supplement Fig 1.

## Results

### ADCC

#### ^51^Cr-Release assays

The CFS patients had lower ADCC capacity per effector cell than that of the unrelated healthy controls. Their family members also had lower ADCC than the controls. ADCC was evaluated using two measurements, CX1:1 and CX-slope. Both measurements are unaffected by CD16A V and F allelic recognition of antibody due to the high amount of antibody bound to the Daudi target cells and the high affinity of obinutuzumab antibody for both allelic forms of CD16A. CX1:1 measures the percentage of ‘target’ cells that are killed at a 1:1 ratio of effector to target cells (E:T) and reflects the fraction of CD16Areceptor positive NK cells that actually display ADCC lytic capacity. (The remaining CD16Apos NK cells may still produce cytokines instead of killing antibody-coated targets.) CX-slope indicates the increase in cells killed as the E:T increases and reflects cellular cooperatively when several lymphocytes attack a single target cell. Representative CX1:1 values and CX-slopes for a CFS patient and a healthy control are indicated in Fig 2A. Fig 2B indicates results calculated by a different method that has been widely used for comparisons, lytic units. Use of lytic units is biased when the slopes differ and will be discussed later. The CX1:1 and CX-slope measurements indicated statistically significant lower values for the CFS patients than for the unrelated healthy controls, see red bars and brackets, Figs 2C and 2D. Notably, CX1:1s and CX-slopes were similar for the CFS patients compared to their unaffected family members. When the data for all the family members were combined, it became evident that all the family members had lower ADCC compared to the unrelated healthy controls (purple bars and brackets, P<0.01). In aggregate, the data contra-indicate low ADCC as a biomarker to diagnose CFS patients, particularly in comparison to their first degree relatives.

A similar trend, of lower ADCC of both CFS patients and family members *vs*. unrelated healthy controls, applied when lytic units were applied for qualitative reference to prior NK studies. The substantial variations in the CX-slopes among donors obviated quantitative comparison of lytic units which require parallel slopes [55]. For example, the lower CX-slope of the CFS patient in Fig 2A resulted in a large lytic unit (requiring so many cells for 50% ADCC that the intersection of the line of cytotoxicity with a y value of 50% ADCC is off the chart). The LU_50_ measurements of the CFS patients were lower compared to family members without CFS (means 98 +/- 195 *vs*. 149 +/- 170 LU_50_/10^6^ ADCC NK effectors, respectively), and lower compared to unrelated healthy controls (mean 187+/- 108 LU_50_/10^6^ ADCC NK effectors). Consequently, due to the increased variations, the P values were statistically insignificant (P = 0.55 for patients *vs*. unrelated healthy controls). We provide the lytic unit calculations to indicate why separate evaluations of CX1:1 and CX-slope are better suited for comparisons of ADCC.

With a disease like ME/CFS that could have several disease subtypes [60, 61], it is possible that the low ADCC applied to only a few of the families and was sufficient to skew the averages for all 5 families. Upon examination, when each family was considered separately, the family-associated lower ADCC (compared to the unrelated healthy controls) applied to each one of the five families (supplement Fig. 1A, 1B, 1C and 1D). The CX1:1 and CX-slopes were lowest for family 10. Also, the CFS patients ran the full gamut from the highest to the lowest values within each family, *e.g*., see family 28 in Figs S1A and S1B for CX1:1 and CX-slopes, respectively. The conclusions from the overall mean values are consistent with the data for each separate family.

It is critical for interpretation of the data that the ADCC of *both* the CFS patients and their unaffected family members was lower than that of unrelated healthy controls. The CX1:1 of the patients was 66% of the CX1:1 of healthy controls, while that of family members without CFS was 82% of the controls. The CX-slopes of the patients was 71% of the controls, while the CX-slopes of the family members without CFS was 78%. The difference between the CFS patients *vs*. the unrelated healthy controls could have been misinterpreted as evidence for low ADCC as a CFS biomarker. However, the ADCC of *both* CFS patients and their family members without CFS was low and there were no statistically significant differences between them. For that reason, inclusion of the family members in the study provided an extremely valuable perspective to eliminate a false biomarker. The coincident observations of lower CX1:1 and lower CX-slope are to be expected: a strong correlation between CX1:1 and CX-slopes applied to all blood donors (supplement Fig 2).

### ROC assessment of biomarker potential

ROC (receiver operating characteristic) plots indicate that low ADCC CX1:1 and low CX-slope have limited potential as biomarkers specific for CFS, particularly when family members without CFS are included as controls. ROC curves represent a means to simultaneously evaluate sensitivity and specificity of a diagnostic test. By design, the areas under the ROC curves (AUC) range from a theoretical perfect value of 1 (for a 100% reliable test for a diagnostic biomarker) to 0.5 which indicates no value as a biomarker. The CFS patients were the true positives for the ROC analyses. For CX1:1, the AUCs comparing positive CFS patients with 3 negative groups of either unrelated healthy controls, all donors without CFS (unrelated healthy and family members without CFS, combined), or family members without CFS were 0.76, 0.72 and 0.69, respectively (Fig 3A). For CX-slope, the AUCs comparing CFS patients with either unrelated healthy controls, all donors without CFS, and family members without CFS were 0.77, 0.69 and 0.45, respectively (Fig 3B). For both ADCC measurements, the ROC analyses followed a similar trend in which the AUC values for unrelated healthy donors were larger than the AUCs for family members without CFS. For the unrelated healthy donors, P<0.05 significance applied to the AUC analyses of the CX-slopes and was close to significant for the CX1:1 measurements (Table 3). However, for both CX1:1 and CX-slopes there were no detectable differences in AUCs between the CFS patients and their unaffected family members. Thus, the ROC measurements indicate that ADCC capacity is an unsuitable biomarker for diagnosis of CFS, particularly when applied to first degree relatives of CFS patients.

**Table 3.** ROC Assessments of Low ADCC as a Biomarker for CFS^a^

| **Table 3. ROC Assessments of Low ADCC as a Biomarker for CFS<sup>a</sup>** |  |  |  |
| --- | --- | --- | --- |
| **True vs. False Test Groups** | **ADCC** | **Area Under Curve** | **P<sup>b</sup>** |
| **CFS patients vs. Family w/o CFS** | CX1:1 | 0.69 | 0.29 |
| **CFS patients vs. All Donors w/o CFS** | CX1:1 | 0.72 | 0.09 |
| **CFS patients vs. Unrelated Healthy Donors** | CX1:1 | 0.76 | **0.06** |
| **CFS patients vs. Family w/o CFS** | CX-slope | 0.45 | 0.92 |
| **CFS patients vs. All Donors w/o CFS** | CX-slope | 0.69 | 0.08 |
| **CFS patients vs. Unrelated Healthy Donors** | CX-slope | 0.77 | **0.02** |

<sup>a</sup>CFS patients met the Fukuda standards. "All donor without CFS" are the family members without CFS and the unrelated healthy controls.
<sup>b</sup>P values by maximum likelihood tests for difference of AUCs from 0.5.

### NK ADCC effector blood cell counts

CD16Apos NK blood cell counts (of the ADCC effector lymphocytes) were lower overall for CFS patients *vs*. unrelated healthy controls but indistinguishable between CFS patients and their family members without CFS. Average CD16Apos NK blood cell counts for the CFS patients were lower (74% of control counts) than that of the unrelated healthy controls, though without statistical significance (Fig 4A). The CD16Apos blood NK cell counts for the family members without CFS were also lower, 78% of the controls. The cell counts of all family members combined was 77% of that of the unrelated healthy controls (purple bars, Fig 4A) and was statistically significant by one-tailed analysis, P <0.05. (In this case, the one-tailed assumption was that the counts would be lower than the unrelated healthy controls). These CD16Apos NK cell counts of patients and their family members without CFS were widely distributed (high and low) within each family (supplement Fig 1C). Thus, the low CD16Apos NK cell counts varied independently of CFS status. Three of four families evaluated (#10, 22, and 28) had low overall CD16A counts while one family (#3) was similar to the unrelated healthy controls. The low effector cell counts had the potential to further decrease ADCC resistance to viral infections in most, but not all, of the CFS families. The antibody panel and flow cytometric gating for the NK CD16Apos ADCC effector blood cell counts are illustrated in Fig 4B.

### Synergistic risk of low ADCC lytic capacity and low effector cell numbers

The combination of low *in vitro* ADCC capacity plus low *ex vivo* effector blood cell counts in individual subjects represents a potential synergistic risk for low *in vivo* ADCC. If an individual were to have low ADCC capacity per cell combined with fewer total ADCC effector cells in their blood, this combination would result in low systemic ADCC capacity. This combination was observed for CFS patients and their family members. First, before querying for combined risks, we assessed if the ADCC cytotoxic capacity and the cell counts varied independently among the donors. The CX1:1s and CX-slopes varied independently from CD16Apos NK effector blood cell counts for all the donors; the R^2^ for linear fits were low (<0.1) and the P values for linearity were insignificant. Therefore, evaluation for synergy is valid. Quadrant analyses were applied to determine if the CFS patients and the CFS family members were over-represented in the low-low quadrants for low CX1:1 combined with low blood effector cell counts (or CX-slope combined with counts). The CFS patients and their family members were 24-fold more likely than unrelated healthy donors to be in the low-low quadrant for CX1:1 and ADCC effector cell counts (Fig 5A, P = 0.02). The CFS patients and their family members were also 12.5-fold more likely than unrelated healthy donors to be in the low-low quadrant for CX-slope and ADCC effector counts (Fig 5B, P<0.05). Only one of the unrelated healthy control donors (indicated by black symbols) was in a low-low quadrant, of Fig 5B. Overall, these two quadrant analyses indicate synergistic familial risks for low *in vivo* ADCC.

**Fig 5.**
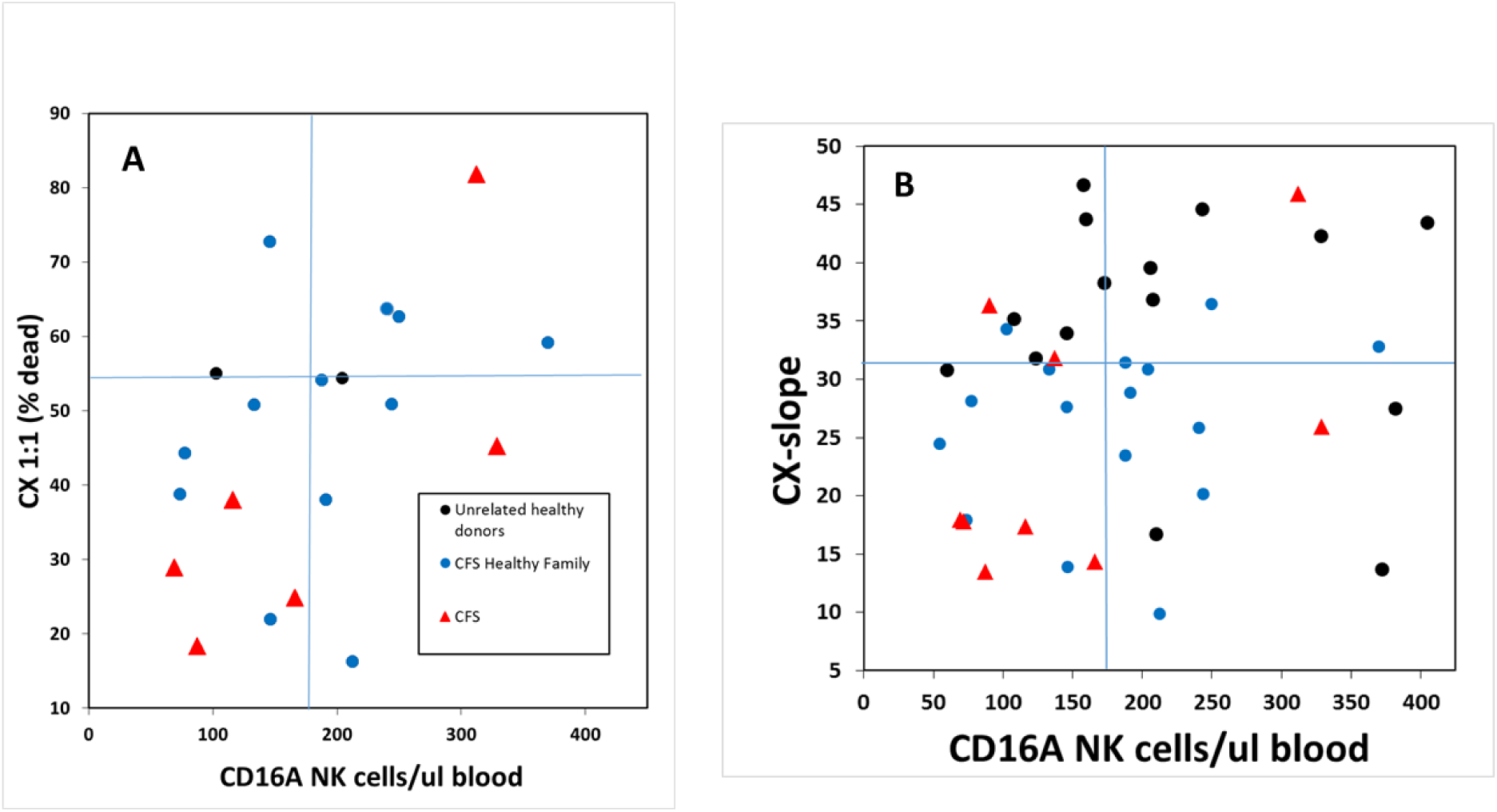
Synergistic risk for low NK cell ADCC and low CD16Apos NK cell counts/µl blood for CFS families compared to unrelated healthy controls. Synergistic risk was assessed by comparing the relative distribution of CFS patients, family members and healthy controls by quadrant analysis to determine if CFS family members were over-represented in the combined low ADCC-low cell number (lower left) quadrant. The data are divided into quadrants based on the median values of CX1:1 (**A**) or CX-slope (**B**) *vs*. blood cell counts for the complete set of donors that included the CFS patients, non-CFS family members, and healthy controls. The ADCC values and the CD16Apos NK cell counts varied independently with P values for linear correlation of 0.27 and 0.15, respectively, statistically non-significant, so that it is appropriate to search for synergy. The CFS patients are indicated with red triangles, the family members without CFS with blue circles and the unrelated healthy controls with black circles. Statistics were applied to assess the relative likelihood of CFS family members *vs*. healthy control appearing in the low-low (lower left) quadrants. **A. Twenty-four fold greater likelihood for the CFS family members to have low CX1:1 combined with low CD16Apos NK cell counts**. Data are from families #3, 10 & 28. For the CX1:1 the median value was 54.4 and for the CD16Apos NK counts the median was 179.6 cells/ul. The 95% confidence limits were 1.7, 330; P= 0.02. **B. Twelve and half fold greater likelihood for the CFS family members to have the low – low combination of low CX-slopes with low CD16Apos NK cell counts. \***P<0.05. For the CX-slope the median value was 30.8 and for the cells/µl the median was 169.1. Data are from families #3, 10, 22 & 28. The 95% confidence limits were 1.1, 143.1; statistically significant at the 5% level.

### *FCGR3A* genetics

*FCGR3A* homozygous genotypes encoding CD16A aa158 F/F represent an additional risk factor for low ADCC because the F/F NK cells require higher amounts of antibody to mediate ADCC than NK cells homozygous or heterozygous with CD16A aa158 V. The CD16A-158F variant has two-fold lower affinity for IgG1&3 than the 158V variant [41]. The F/F genotype is associated with lower *in vitro* ADCC at low antibody concentrations than the F/V and V/V genotypes [57, 62]. The frequency of the F/F genotype was 91% for the CFS patients and 82% for the family members without CFS, while the unrelated healthy controls were 18.8% F/F (Table 4 and pedigrees, Fig 1). The Utah population that includes the subjects of this study is of non-Finnish northern European descent [63, 64]. The F/F frequency for such a European group (with 919 sampled) is 41.9% [65]. Thus, the F/F frequencies of the CFS patients and family members are higher than would be predicted while the F/F frequencies of the controls are lower than would be predicted.

**Table 4.**
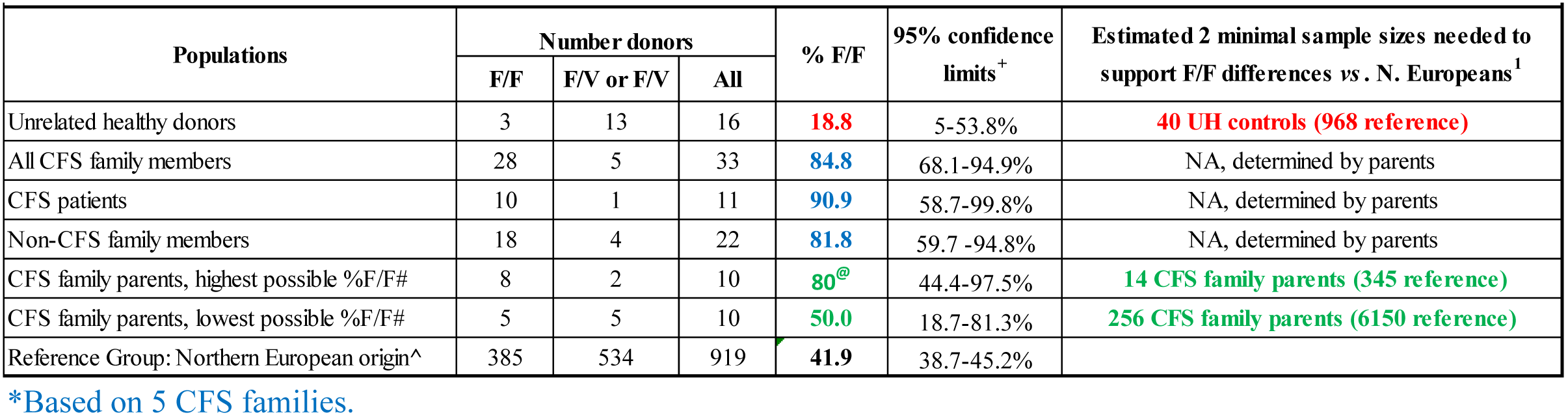
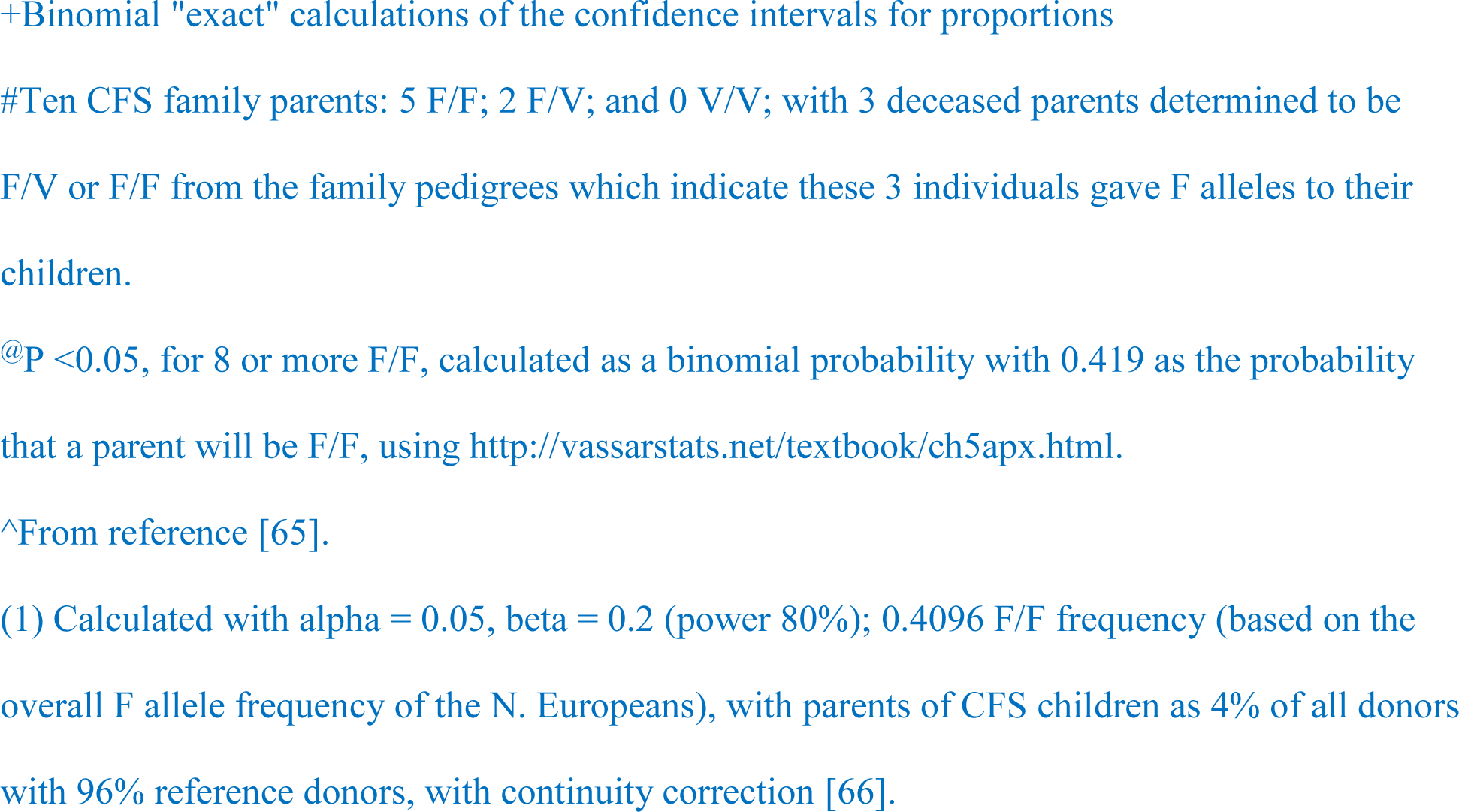
Frequencies of CD16A F/F homozygosity*

| Table 4. Frequencies of CD16A F/F homozygosity* |  |  |  |  |  |  |
| --- | --- | --- | --- | --- | --- | --- |
| Populations | Number donors |  |  | % F/F | 95% confidence limits<sup>+</sup> | Estimated 2 minimal sample sizes needed to support F/F differences vs. N. Europeans<sup>1</sup> |
|  | F/F | F/V or F/V | All |  |  |  |
| Unrelated healthy donors | 3 | 13 | 16 | **18.8** | 5-53.8% | **40 UH controls (968 reference)** |
| All CFS family members | 28 | 5 | 33 | **84.8** | 68.1-94.9% | NA, determined by parents |
| CFS patients | 10 | 1 | 11 | **90.9** | 58.7-99.8% | NA, determined by parents |
| Non-CFS family members | 18 | 4 | 22 | **81.8** | 59.7-94.8% | NA, determined by parents |
| CFS family parents, highest possible %F/F# | 8 | 2 | 10 | **80<sup>@</sup>** | 44.4-97.5% | **14 CFS family parents (345 reference)** |
| CFS family parents, lowest possible %F/F# | 5 | 5 | 10 | **50.0** | 18.7-81.3% | **256 CFS family parents (6150 reference)** |
| Reference Group: Northern European origin<sup>^</sup> | 385 | 534 | 919 | **41.9** | 38.7-45.2% |  |

\*Based on 5 CFS families.
+Binomial "exact" calculations of the confidence intervals for proportions #Ten CFS family parents: 5 F/F; 2 F/V; and 0 V/V; with 3 deceased parents determined to be F/V or F/F from the family pedigrees which indicate these 3 individuals gave F alleles to their children. @P <0.05, for 8 or more F/F, calculated as a binomial probability with 0.419 as the probability that a parent will be F/F, using http://vassarstats.net/textbook/ch5apx.html. ^From reference [65]. (1) Calculated with $\alpha = 0.05$ , $\beta = 0.2$ (power 80%); 0.4096 F/F frequency (based on the overall F allele frequency of the N. Europeans), with parents of CFS children as 4% of all donors with 96% reference donors, with continuity correction [66].

An initial response would be to compare all the family members with their genealogical group or with the healthy controls of the study, but this comparison fails to consider that F/F by F/F parents will produce only F/F offspring and that the number of these progeny is irrelevant. The appropriate analysis is to first examine the pedigrees for F/F x F/F parental pairs (the first generation) and use the F/F frequency of the 10 parents for statistical analyses. The appropriate issue for this study is the significance of the F/F frequencies of these parents *vs*. the predicted F/F incidence of the general population.

#### Two estimates (for the highest and lowest possible parental F/F frequencies) were used to assess the potential statistical significance of the parental F/F homozygosity

The 7 available parental genotypes are 5 F/F and 2 F/V. The other 3 of the 10 parents were deceased; their genotypes, as either F/F or F/V, were determined from the genotypes inherited by their progeny. (F/F progeny had to inherit an F allele from the deceased parent who had to have at least one F allele, see pedigrees Fig 1.) The highest possible F/F parental frequency is 8 F/F & 2 F/V parents (80% F/F) and is likely based on their 8 progeny who were all F/F, see Table 4, line 6. The lowest estimate is 5 F/F & 5 F/V parents (50% total F/F); see Table 3, line 7. The high estimate of 80% F/F parents was statistically significant compared to the 919 northern European controls (P = 0.02) while the low estimate was insignificant (P = 0.42). These calculations are based on all 5 families and, because of the uncertainty of the genotypes of the deceased, are only indications of potential significance.

### Sample sizes required for better assessment of CD16A F/F in CFS

To guide future researchers to address the risk of CD16A F/F homozygosity for CFS in other geographic populations, we estimated the minimal sample sizes of CFS family parents and controls needed to support F/F frequencies as a risk factor. We present a model scenario that assumes an incidence of CD16A F/F homozygosity of 0.4096 based on the F allele frequency of the northern Europeans, a prediction of 80% F/F CFS parental homozygosity associated with CFS, and that the researcher will be able to amass a sample population with 4% of the donors who are parents with a CFS child. The model includes consideration of type 2 statistical errors. The last column of Table 4 provides the predictions for the minimal numbers of donors needed in the CFS parental and control groups. Fourteen parents with 345 unrelated controls would be needed for the 80% hypothetical F/F parental frequency, suggesting that examination of additional populations would be feasible. A larger number (256) of parents would be required to support 50% F/F parental frequencies with 6150 controls; these numbers are currently impractical and underscore the problems associated with pilot clinical research studies (such as this report) where limited sample sizes hamper interpretations [67, 68].

### Recognition of antibodies bound to ADCC target cells

EC50s are the effective concentrations of antibody needed to support 50% of the maximal ADCC that a fixed number of lymphocytes can mediate and indicate the ability of NK cells to recognize antibody bound to target cells. EC50s are influenced by CD16A alleles. CD16A F/F donors have only the lower affinity receptors for the Fc of IgG antibodies in contrast to F/V and V/V donors with higher affinity V allele [56, 69]. CD16A F/F NK cells usually require higher concentrations of antibody to support ADCC. High EC50s were required to support ADCC by the CFS family members and represent an immune risk factor. The EC50s of the F/F CFS patients and their F/F family members without CFS were similar, with average EC50s of 4.0 and 3.3 ng/ml obinutuzumab, respectively, much higher than the unrelated healthy control average EC50 of 0.33 ng/ml (Fig 6, P<0.05). These EC50 differences are consistent with differences between F/F CFS families and the control donors with V alleles. A potentially noteworthy observation is that the EC50s values of the family members were bimodal and remarkably high for some members. Without the unusually high values, the average EC50s for CFS patients or their family members (0.8 & 0.9 ng/ml, respectively) were similar to the three unrelated healthy F/F controls (0.6 ng/ml). As expected, these EC50s are still significantly higher than the unrelated controls with V alleles. The previously reported higher antibody concentrations needed to support ADCC by cells with CD16A F/F genotypes are consistent with the CD16A F/F participants in this study. The ultimate significance to CFS is directly linked to the unresolved issue of whether CFS patients actually have high frequencies of CD16A F/F genotypes.

**Fig 6.**
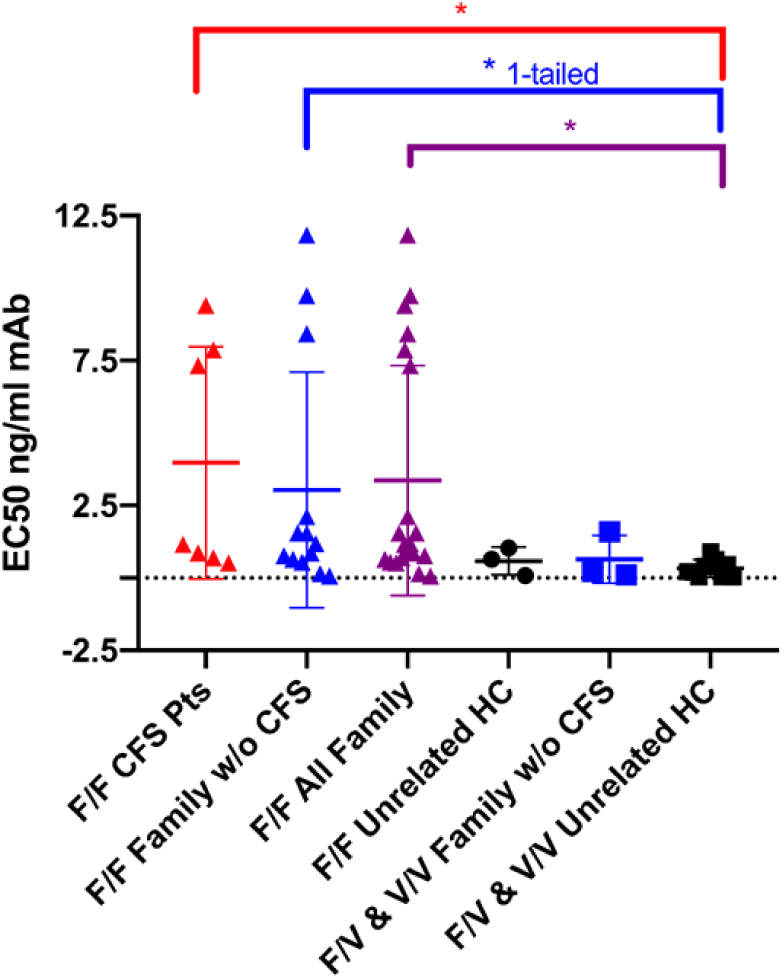
Antibody EC50s. The EC50s of CFS patients and family members without CFS were higher than those of unrelated healthy donors. These EC50s are attributable to the CD16A genotypes in which the V allele encodes a higher affinity receptor for the IgG1 antibody used. The data are from families 3, 20 and 22. The CFS patients and their family members without CFS who were homozygous F/F had EC50s that were higher than the EC50s of the unrelated healthy controls (who had F/V & V/V genotypes). The EC50s of the CFS patients and family members were bimodal; when values >3 ng/ml were excluded the EC50s were still greater than the unrelated controls (P<0.05, not illustrated). The data are consistent with previous reports that ADCC by NK cells from F/F donors requires higher concentrations of antibody than from donors with the V allele. The numbers of F/F healthy controls (three) and F/V & V/V CFS patients (three) were too low to support statistical comparisons of groups with common genotypes; nonetheless, the average EC50s of CFS F/F family members (3.7 ng/ml) were higher than the F/F unrelated controls (0.6 ng/ml, P= 0.25). Red symbols indicate CFS patients, blue symbols family members without CFS, purple symbols all CFS family members and black symbols the unrelated healthy donors. The bracketed comparisons with asterisks had P values <0.05.

## Discussion

### Major findings

This pilot report presents four major findings. (1) *ADCC of CFS patients was lower than that of unrelated healthy controls but was similar to the ADCC of their family members without CFS*. The family members, both CFS patients and non-CFS, had ∼75% of cytotoxic capacity compared to the unrelated healthy controls. (2) *The ADCC effector cell blood counts were lower for all family members vs. unrelated healthy controls*. (3) *When low ADCC and low ADCC effector cell numbers are considered as synergistic risks, there was a 12 fold or greater risk for CFS family member*s. (4) *Most members of the CFS-affected families had low affinity CD16A F/F antibody receptors*. Here we provide perspective for each finding, then discuss other immune parameters that may affect the ADCC of the CFS patients, and make suggestions for future characterization of ADCC in CFS disease.

Even though this pilot report involved only 11 CFS patients and 5 families, the similarity of ADCC between CFS patients and their family members without CFS clearly indicates lack of potential for low ADCC as a biomarker for CFS. The lower CX1:1 of CFS patients *vs*. unrelated healthy controls could have been mistaken as CFS disease-associated; however, the similarity between the patients and their family members without CFS indicated that CX1:1 lacks specificity as a biomarker for CFS. The same similarity was found for the CX-slope determinations. First degree family members, with common genetic backgrounds and common environmental exposures, improve the chances that disease-specific genes or altered physiology will stand out clearly. As a result, this study is deeply indebted to the many family members who participated. The significant differences that are reported here between NK cell ADCC activities of the CFS patients and unrelated controls and an absence of significant differences between the patients and their unaffected family members were also noted in the first CFS family report of NK activity by Dr. Paul Levine *et al*. [7]. The Levine study monitored antibody-independent NK killing of K562 cells. They reported a lack of statistical difference in NK activities (P=0.38) for 8 family members with CFS compared to 12 unaffected family members. The pattern of lowest NK cell activity of CFS patients, intermediate activity for first degree family members and highest activity for unrelated controls was observed in the Levine report as well as for ADCC in the current report.

### Value of family members without disease for biomarker evaluation

Inclusion of family members without disease offers advantages for the screening of biomarkers specific for diseases. Comparison of patients *vs*. family members without the specific disease offers researchers the ability to “cut to the chase” in pilot studies. As shown in this study, the family members provided an early warning for biomarker evaluation. The family members without CFS had a much higher false positive rate as compared to the unrelated healthy control subjects. Inclusion of family members can also identify disease-independent differences between family members and the unrelated healthy controls, as we observed in this study. The ROC analyses for CX1:1 and CX-slope provide strong support for separate assessments of an unaffected family member control group as well as a group of unrelated healthy controls who are matched with the patients for sex, age, and race. The ROCs indicate some potential for low ADCC as a CFS biomarker for persons unrelated to CFS patients and unsuitability of low ADCC as a test for CFS for persons who are first degree family members of CFS patients. Similar ROC AUC values were reported for evaluation of (low) NK cell-mediated natural cytotoxicity to K562 cells as a biomarker for CFS patients when they were compared *vs*. unrelated healthy controls [31]. For future studies, we highly recommend inclusion of family members without CFS as a second control group.

### Limitation of this report

This report has the limitations of only one race, donors from a single geographic location, and a CFS patient population with moderate disease rather than extreme completely bed-ridden disease. A study with the same race but a Scandinavian location, with 48 unrelated CFS patients, came to the same conclusion as this report: ADCC is unsuitable as a biomarker for CFS [38]. The Scandinavian study used different methods to assay ADCC and, in contrast with this Utah-based study, found no significant differences between CFS patients and controls. The two different geographic locations (USA and Scandinavia) increase support for the unsuitability of ADCC as a CFS biomarker. The effects of low ADCC in CFS for other races and in extreme CFS disease are still unanswered.

### ADCC as a risk factor in CFS

When low ADCC lytic capacity and low effector cells were considered as synergistic risk factors predisposing for CFS, there was a greater risk for all the CFS family members. The reductions affecting ADCC (cytotoxic capacity and numbers of ADCC effector cells) that were reported here were modest and unlikely alone to cause disease. As observed with type 1A autoimmune diabetes [70], multiple risk factors that are immunological in nature can predispose individuals to diseases that may be triggered by infections.

The high incidence of the CD16A *FCGR3A* F/F genotype in the CFS families indicates that genetics, as well as environmental factors such as viral infections, may contribute an additional risk of CFS. The F/F incidence was strikingly high, but the small sample size of the parents (and incomplete genotypes for three deceased parents) impacted the statistical analyses, leaving F/F homozygosity as a CFS risk factor an unresolved issue. Since the F allele is common in the human population, even if F/F status is a risk factor for CFS, it is likely that there are other genetic risk factors. Twin studies, first published by Dr. Deborah Buchwald *et al*. [71], initiated evidence for such genetic risks. Pair-wise concordance rates of CFS in monozygotic twins range between 8%-38% in different reports, and are greater for monozygotic compared to dizygotic twins and compared to other family members [25, 26, 71-74]. It is relevant to ADCC that pairs of monozygotic twins (without CFS) have similar frequencies of CD16Apos NK cells and CD16Apos NK subsets in contrast to the differences observed among unrelated individuals [75]. This twin study of NK cells also documented differences between pairs of monozygotic twins for their NK cell subsets, indicating that non-genetic influences profoundly affect diverse NK populations. Additional studies, both functional and genetic, of NK cell immunity in CFS patients *vs*. family members without CFS, have the potential to reveal CFS-associated alterations in immunity and to find disease-associated biomarkers.

The CFS patients may have critical combinations of immunological risks that can lead to poor responses to a variety of infectious challenges. The cause(s) of CFS – genetic, viral, or other – continue to confound the best of scientists [76]. If multiple risk genes are encoded on different chromosomes, one would predict that, due to independent chromosomal segregation at meiosis, a CFS patient could inherit a critical number of risk alleles, while their non-CFS siblings could have some, but not the entire set, of risk alleles. This scenario could affect ADCC because several genes other than *FCGR3A* affect ADCC activity. For example, inheritance of inhibitory NK cell KIRs together with alleles of MHC class I proteins that bind the KIRs will reduce ADCC [77]. Inheritance of G1m IgG allotypes with less affinity for CD16A will reduce ADCC efficacy [78]. Homozygous lower affinity G1m_17_ antibody in combination with CD16A F/F NK cells resulted in lower *in vitro* ADCC activity toward cells infected with the chronic virus *Herpes simplex 1* compared to G1m_3/3_ anti-viral antibody with CD16A V/V NK cells [79]. Antibody fucosylation varies [80] and only non-fucosylated antibodies support ADCC [81]. The cytokine TGFβ is elevated in patients with CFS [21], and TGFβ promotes loss of CD16A from NK cells [82]. Regulatory factors, such as microRNAs that are elevated in the NK cells of CFS patients [83], may influence ADCC. These examples indicate that many differences among people, including hereditary differences, are candidates to affect ADCC and to be CFS risk factors in addition to *FCGR3A* F/F homozygosity. Furthermore, the fatigue induced by chronic viruses that are controlled by ADCC, such as after initial infection with Epstein Barr Virus to cause mononucleosis, resembles symptoms of CFS [84]. Fatigue associated with viral infections provides appeal for the hypothesis of compromised ADCC toward chronic viruses in the pathogenesis in CFS. However, increased fatigue associated with post-exertional malaise occurs without evidence for viral reactivation [85] and indicates just how difficult it is to establish links between chronic infection and CFS disease. Recently, there has been interest in intestinal infections in CFS [86] which, if conclusive, would indicate that previous researchers may have had the right ideas and been looking in the wrong places.

### Implications for future research

Continuation of research on ADCC in CFS presents opportunities to benefit the patients. It is tempting to propose a mechanistic linkage in which low ADCC sub-optimally controls chronic herpes viruses that could reactivate to promote the fatigue and neurological symptoms of CFS patients -- even though viral etiology remains unproven. Comparison of CFS patients and their family members without CFS may reveal additional genetic variants other than *FCGR3A* alleles that may affect ADCC. Unfavorable IgG Gm1 allotypes, low levels of the IgG1 & IgG3 antibodies that support ADCC (reported for some CFS patients [87]), and high antibody fucosylation may help explain why some CFS patients benefit from antibodies from unrelated donors that are included within intravenous immunoglobulin (IVIG) therapy [88-91]. Identification of those CFS patients with low CX1:1 capacities may indicate those patients most likely to respond to immunomodulatory therapies such as poly(I:C) therapy [92, 93] that promotes NK cell cytotoxic functions [94]. Discovery of the inflammatory sites of production of TGFβ in CFS patients is important because localization of TGFβ to reservoirs of chronic viruses may provide clues to key immune responses that are undetectable when blood is sampled. In summary, this report adds support for a role for partial immunodeficiencies in CFS pathology.

## Data Availability

Data are available upon request.

## Acknowledgments

We are deeply grateful to the patients and their families who participated in this study and to the leadership and staff at the Bateman Horne Center who made this research study possible. Suzanne D. Vernon, Ph.D., at the Bateman Horne Center contributed valuable guidance throughout the project. We thank Roche Pharmaceutical Research & Early Development for the non-fucosylated type 2 anti-CD20 monoclonal antibody obinutuzumab (Gazyva®, GA101 G2). DNA was isolated by Ms. Laura Meadows at UNR and the *FCGR3A* aa158 alleles sequenced at the NIH by Stephen K. Anderson, Ph.D. We are grateful for the advice of Lynn B. Jorde, Ph.D., University of Utah School of Medicine, for help with interpretation of the *FCGR3A* genetic data. We thank Parker Hoshizaki, Terry Woodin, Ph.D., and Dr. Stephen K. Anderson for manuscript editing. Dr. Redelman died before the submission of the final version of this manuscript. Dr. Hudig accepts responsibility for the integrity and validity of the flow cytometric data he collected and analyzed.

## Author Contributions

Contributions are arranged alphabetically.

**Conceptualization:** Dorothy Hudig, Doug D. Redelman **Data Curation:** Michael J. Guglielmo, Dorothy Hudig

**Formal Analysis:** Michael J. Guglielmo, Dorothy Hudig, Julie Smith-Gagen, Doug D. Redelman, Alexander P. Sung

**Funding Acquisition:** Dorothy Hudig

**Investigation:** Michael J. Guglielmo, Dorothy Hudig, Doug D. Redelman, Alexander P. Sung, Jennifer J-J Tang

**Methodology:** Dorothy Hudig, Doug D. Redelman

**Project Administration:** Dorothy Hudig, Lucinda Bateman

**Resources:** Lucinda Bateman

**Supervision:** Dorothy Hudig

**Validation:** Dorothy Hudig

**Visualization:** Michael J. Guglielmo, Dorothy Hudig, Alexander P. Sung, Jennifer J-J Tang

**Writing - original draft:** Alexander P. Sung

**Writing– review & editing:** Lucinda Bateman, Dorothy Hudig, Michael J. Guglielmo, Alexander P. Sung. Julie Smith-Gagen

## Abbreviations

aa: amino acid
ADCC: antibody-dependent cell-mediated cytotoxicity
CD16A: cluster of differentiation protein 16A, the IgG Fc-receptor of NK cells
CFS: chronic fatigue syndrome
CX1:1: the % cells killed at a 1:1 ratio of CD16Apositive NK cells to ^51^Cr-labeled Daudi cells with pre-bound obinutuzumab anti-CD20 mAb
CX-slope: cytotoxic slope as the effector cells are increased
E:T: effector to target cell ratio
EC50: the effective concentration of antibody needed for 50% of maximal ADCC
FcR: cellular receptor for the Fc region of immunoglobulin (antibody)
*FCGR3A*: the gene encoding CD16A, the IgG FcR of NK cells
KIR: killer cell immunoglobulin-like receptor
ME: myalgic encephalomyelitis
NK: natural killer lymphocyte
LU_50_: lytic unit_50_, the number of effector cells needed to kill 50% of the ‘target’ cells
PBMC: peripheral blood mononuclear cells
ROC: receiver-operating characteristic plot
UHC: unrelated healthy control subject

